# EFFORT TO ACHIEVE QUALITY JOB THROUGH INCREASING NURSE PERFORMANCE MOTIVATION

**DOI:** 10.1101/2022.09.03.22279569

**Authors:** Achmad Masfi, Moses Glorino Rumambo Pandin

## Abstract

**Objective:** The era of globalization is a challenge for health companies to keep abreast of trends and issues in the health sector. The challenges of health companies are not only external factors, Human Resources is one of the challenges that must be solved by health companies, especially the motivation of nurses.

**Method:** The method in this research is Literature Review. The purpose of this research is to improve company performance through employee motivation. The search method uses the Prisma 2020 chart, the literature search for the authenticity of this research is carried out with a literature review using the keywords “Performance Motivation” AND “Quality Jobs”“ from 2020 to 2022. The search results obtained 8,430 articles but only 25 articles that match the criteria study. Articles obtained from Scopus, ScienceDirect, ProQuest and PubMed, google scholar

**Findings:** The results of 15 journal articles related to Performance Motivation, show that motivation affects employee performance, both motivation as an independent variable, dependent variable or intermediary variable, all show that there is a very strong relationship between motivation and employee performance in general. Based on previous research, there is a positive and significant influence between work motivation and employee performance

**Conclusion:** Based on the results of the Literature Review, it was found that there is a positive and strong relationship between Performance Motivation and Work Quality. As an effort to improve the quality of work, it is necessary to meet the variables that affect work motivation such as psychological motivation, safety motivation, social motivation, reward motivation, and actualization motivation, leadership, etc.

## Introduction

The era of globalization is a challenge for companies to keep abreast of trends and competitiveness. The company’s challenges are not only external factors, but internal factors. One of the internal factors that become a challenge is Human Resources. Therefore, to face the challenges of globalization, there needs to be efforts to develop quality human resources (HR), ready to compete with global competencies, proven scientific foundations, mature in mature human resource skills [1]

Research studies related to Human Resources are often associated with the quality of performance and also performance motivation. Research on motivation is often associated with the behavior and attitudes of nurses which will certainly have an impact on the quality of nurse performance. Several studies on motivation are always associated with financial elements, most of the leaders of health companies do not know about efforts to increase motivation, they only think to increase motivation by increasing financially, even though finance is not necessarily able to increase nurse motivation. The importance of motivation in the world of health, especially nurses, has a very large role in developing and completing nurses’ duties in a timely manner and with good quality. The manager level can be a motivator for nurses, while nurses must be a motivator for their patients. Copy that high motivation must also be intertwined between one nurse and another nurse [2]

Human resource management must be carried out effectively, efficiently and professionally so as to create a comfortable working atmosphere. A comfortable atmosphere will create a good relationship between nurses, work harmony and help achieve the health company’s achievements. Harmony, and cohesiveness between nurses will create a productive and professional performance. The development of a company is not based on capital or finance, but the success of a company is determined by how it manages human resources [3]

Improving the quality of work, especially in health companies is the goal of The Sustainable Development Goals (SDGs). One of the Sustainable Development Goals (SDGs) goals is SDG’S 8 Decent work and economic growth there is “Quality jobs” which means that the quality of work is one of the SDG’s 2020 Goals, so in this paper it supports the SDG’s goal in improving quality jobs through employee motivation. [4]

## Method

This study uses the Literature Review method. The purpose of this study is to improve company performance through employee motivation. The search method uses the Prisma 2020 chart, the literature search for the authenticity of this research is carried out with a literature review using the keywords “Performance Motivation” AND “Quality Jobs”“ from 2020 to 2022. The search results obtained 8,430 articles but only 25 articles that match the criteria study. Articles obtained from Scopus, ScienceDirect, ProQuest and PubMed, google scholar

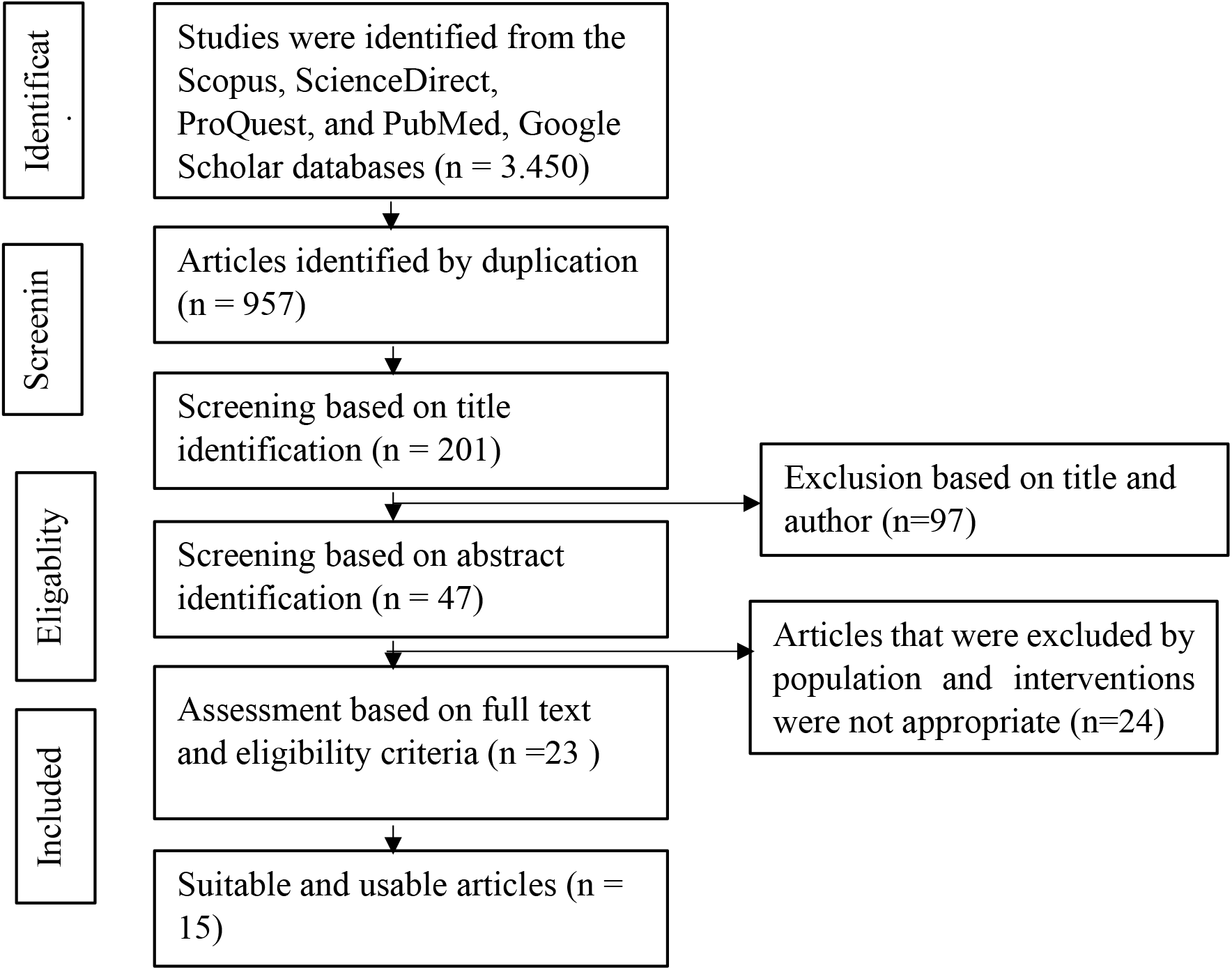
Gambar Pencarian Literatur Menggunakan Prisma 2020 Upaya Dalam Mencapai Pekerjaan Berkualitas Melalui Peningkatan Motivasi Kinerja

## Results

The results of the Literature Review review of 15 journals found using the Prisma 2020 method, starting with the population search process, namely Performance Motivation and Quality Jobs, then carrying out an exclusion process in the year the journal was published, namely 2020 - 2021 as a form of credibility for taking the library in this literature review.The 15 journals are as follows:

**Table 1.1.**
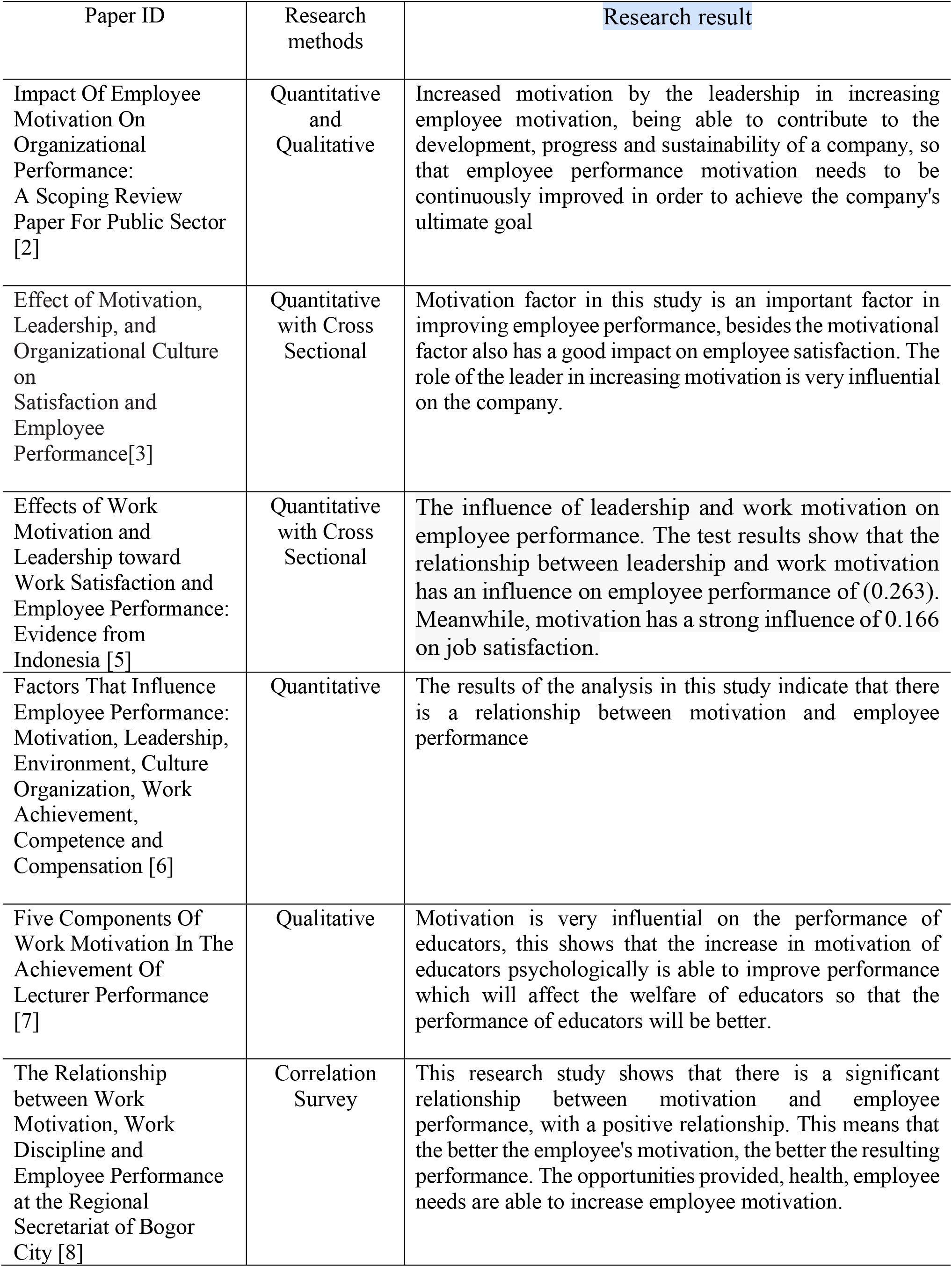
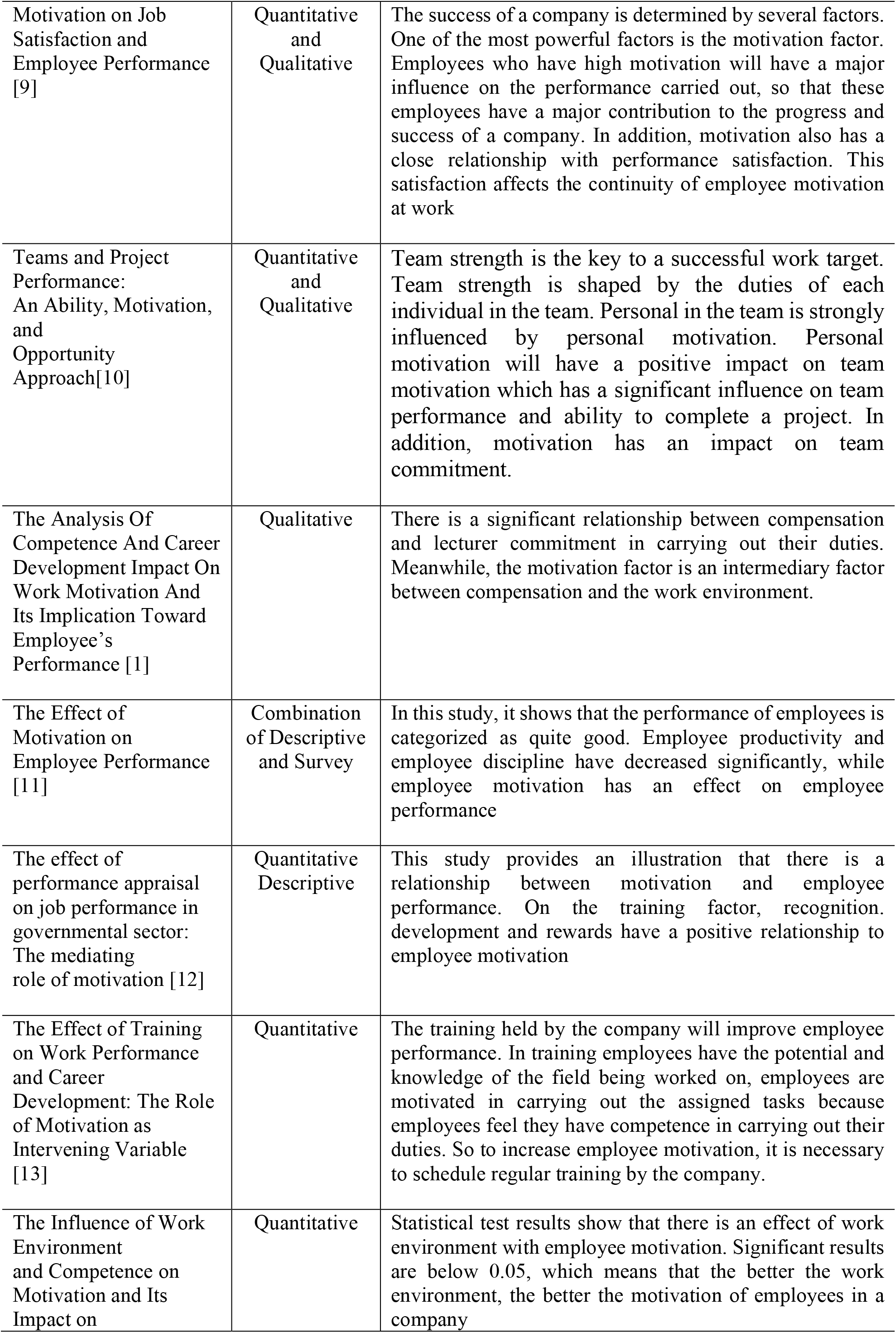
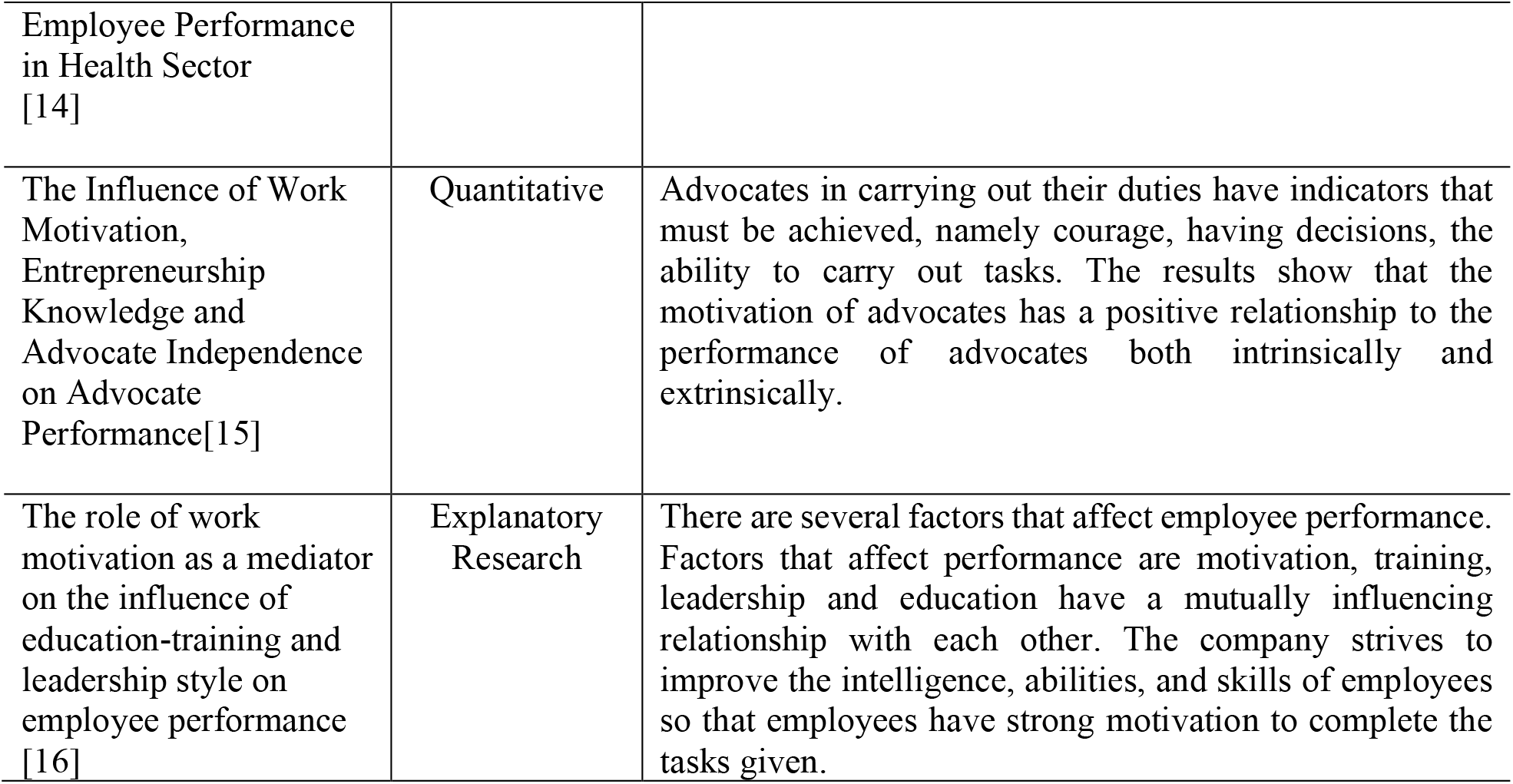
Results of Review of Journal Articles

## Discussion

The results of 15 journal articles related to Performance Motivation, show that motivation affects employee performance in general. Motivation is the main factor in determining employee performance, especially in health companies. The work motivation of nurses will affect the performance and quality of work given to nurses. The higher the motivation of nurses, the higher the performance that will be produced, the better the quality of work provided. This will affect the health company’s target in achieving the goals that have been determined in the company’s vision and mission. Based on the results of the study, employee motivation in general has a very positive influence on employee performance.[17]. In the health sector, it also shows that there is a positive relationship between nurse motivation and nurse performance, nurses who have good motivation, are easy to complete work, are easy to be given responsibility, are easy to be directed so that these nurses have better abilities and experience than nurses who have low motivation.

Motivation is a strong drive that comes from oneself. Motivation is also a way and effort to give one’s enthusiasm and confidence towards a desire to be achieved. There are several factors that will affect motivation, namely, perceptions, needs and desires achieved. Motivation will provide ways and efforts that are more effective and efficient to achieve the desired goals. In addition, motivation also affects other factors within a group or company, such as the work environment, work culture, interactions between one employee and another, commitment and achievement strategies within a company. While performance is the result of a person in carrying out a task given by the leader. In addition to performance is influenced by motivation, performance is also influenced by skills, education, abilities, experience, enthusiasm and time in completing tasks. Performance has quality and quantity as indicators of success in carrying out the assigned tasks. In terms of quantity, the performance is carried out on time, systematically and with alacrity in carrying out the task, while in terms of work quality is the work produced in accordance with the targets, vision and mission determined by the company. In addition, performance is also strongly influenced by intrinsic and extrinsic factors in employees [18].

Motivation and performance variables also have a close relationship with other variables, namely salary, training, work environment, rewards, leadership and performance satisfaction. Motivation has a big role in increasing the satisfaction of health workers or nurses. The satisfaction of the nurse’s performance will be reflected in the implementation of the tasks given, the satisfaction of the nurse’s performance affects the professionalism of the nurse’s performance in behaving and behaving. Nurses who have good work motivation will have a big impact on the health company where they work. The impact of nurses’ motivation on the company is an increase in community visits to obtain health services, a great appreciation from the community in the form of public trust, financial benefits, company development and improvement of quality human resources. Quality human resources will increase good social relations between nurses and other nurses [3]

Efforts that can be made to improve work quality are none other than increasing employee motivation. Meanwhile, to increase employee motivation, it is necessary to increase the variables that affect motivation. According to research conducted by [7], namely psychological motivation, safety motivation, social motivation, reward motivation, and actualization motivation. psychological motivation, described as the fulfillment of welfare for employees. Safety motivation is described as security and comfort as well as employee guarantees in carrying out the tasks at hand

Social motivation, described as a light task load, in accordance with the portion of ability, education and situation. In addition, it is also described as cooperation between one employee and another such as the creation of a comfortable work culture. Reward motivation, described as a form of appreciation for employees, either in the form of material or non-material, this will trigger the enthusiasm of employees in doing a good job. Actualization motivation is described in the form of self-confidence, capacity building, training, achievement and education etc. [7]

Employees who have high motivation will complete the job well, in accordance with what is desired by the company. The work is completed on time, according to the target, and is measured and can be developed for the sustainability of the company. This shows the quality of a work is very good. Indicators of the quality of work are seen from the time, target, speed and accuracy in carrying out the tasks being carried out. The hope of this research is that in improving the quality of work employees need to consider increasing work motivation so that job satisfaction will be achieved for both employees and companies, which will then affect the products to be produced. So that the SDG’s target to improve the quality of work can be achieved as a form of the country’s development process.

## Conclusion

There is a positive and strong relationship between Performance Motivation and Work Quality. As an effort to improve the quality of work, it is necessary to meet the variables that affect work motivation such as psychological motivation, safety motivation, social motivation, reward motivation, and actualization motivation, leadership, etc.

### Limitations

This study has several limitations, such as some articles do not mention the theory or framework that underlies the research and some articles also do not explain the research clearly the instruments that are used, and the secondary results.

### Implementation of Findings in Nursing Practice

This finding supports and becomes a reference for management nursing, and it is also used to improve employee performance, both employees in general, and employees in the hospital, especially the performance of nurses.

## Data Availability

All data produced in the present study are available upon reasonable request to the authors
All data produced in the present work are contained in the manuscript
All data produced are available online at scopus, google scholar, etc

## Daftar Pustaka

[1] H. Nuriman, “The Analysis Of Competence And Career Development Impact On Work Motivation And Its Implication Toward Employee ‘ s Performance,” J. Mhs. Ekon. Bisnis, vol. 1, no. 1, pp. 10–17, 2021.

[2] S. Kalogiannidis, “Impact of Employee Motivation on Organizational Performance.,” Int. J. Adv. Res., vol. 7, no. 10, pp. 166–172, 2021, doi: 10.21474/ijar01/9818.

[3] M. Paais and J. R. Pattiruhu, “Effect of Motivation, Leadership, and Organizational Culture on Satisfaction and Employee Performance,” J. Asian Financ. Econ. Bus., vol. 7, no. 8, pp. 577–588, 2020, doi: 10.13106/JAFEB.2020.VOL7.NO8.577.

[4] ONU, “Sustainable Development Goals: Guidelines for the Use of the SDG,” United Nations Dep. Glob. Commun., vol. 17 SDG’s, no. May, pp. 1–68, 2020, [Online]. Available: https://www.un.org/sustainabledevelopment/news/communications-material/

[5] I. Pancasila, S. Haryono, and B. A. Sulistyo, “Effects of work motivation and leadership toward work satisfaction and employee performance: Evidence from Indonesia,” J. Asian Financ. Econ. Bus., vol. 7, no. 6, pp. 387–397, 2020, doi: 10.13106/jafeb.2020.vol7.no6.387.

[6] P. T. Nguyen, A. Yandi, and M. R. Mahaputra, “Factors That Influence Employee Performance: Motivation, Leadership, Environment, Culture Organization, Work Achievement, Competence and Compensation (A STUDY OF HUMAN RESOURCE MANAGEMENT LITERATURE STUDIES),” Artic. Inf., vol. 1, no. 4, pp. 645–662, 2020, doi: 10.31933/DIJDBM.

[7] Wahyudi, “FIVE COMPONENTS OF WORK MOTIVATION IN THE,” Sci. J. Reflect., vol. 5, no. 2, pp. 466–473, 2022.

[8] Y. Maryani, M. Entang, and M. Tukiran, “The Relationship between Work Motivation, Work Discipline and Employee Performance at the Regional Secretariat of Bogor City (disiplin kerja),” Int. J. Soc. Manag. Stud., vol. 02 No. 02, no. 02, pp. 1–16, 2021, [Online]. Available: https://ijosmas.org/index.php/ijosmas/article/view/14

[9] da C. Carvalho Adelina, I. G. Riana, and A. D. C. Soares, “Motivation On Job Satisfaction And Employee Performance,” Int. Res. J. Manag. IT Soc. Sci., vol. 7, no. 5, pp. 13–23, 2020.

[10] À. Dasí, T. Pedersen, L. L. Barakat, and T. R. Alves, “Teams and Project Performance: An Ability, Motivation, and Opportunity Approach,” Proj. Manag. J., vol. 52, no. 1, pp. 75–89, 2021, doi: 10.1177/8756972820953958.

[11] Y. Kuswati, “The Effect of Motivation on Employee Performance,” Budapest Int. Res. Critics Inst. Humanit. Soc. Sci., vol. 3, no. 2, pp. 995–1002, 2020, doi: 10.33258/birci.v3i2.928.

[12] Y. Al-Jedaia and A. Mehrez, “The effect of performance appraisal on job performance in governmental sector: The mediating role of motivation,” Manag. Sci. Lett., vol. 10, no. 9, pp. 2077–2088, 2020, doi: 10.5267/j.msl.2020.2.003.

[13] D. R. Niati, Z. M. E. Siregar, and Y. Prayoga, “The Effect of Training on Work Performance and Career Development: The Role of Motivation as Intervening Variable,” Budapest Int. Res. Critics Inst. Humanit. Soc. Sci., vol. 4, no. 2, pp. 2385–2393, 2021, doi: 10.33258/birci.v4i2.1940.

[14] R. D. Parashakti, M. Fahlevi, M. Ekhsan, and A. Hadinata, “The Influence of Work Environment and Competence on Motivation and Its Impact on Employee Performance in Health Sector,” vol. 135, no. Aicmbs 2019, pp. 259–267, 2020, doi: 10.2991/aebmr.k.200410.040.

[15] S. Prayetno and H. Ali, “The influence of work motivation, entrepreneurship knowledge and advocate independence on advocate performance,” Int. J. Innov. Creat. Chang., vol. 12, no. 3, pp. 147–164, 2020.

[16] L. Fonseca Da Costa Guterresa, Armanu, and Rofiaty, “The role of work motivation as a mediator on the influence of education-training and leadership style on employee performance,” Manag. Sci. Lett., vol. 10, no. 7, pp. 1497–1504, 2020, doi: 10.5267/j.msl.2019.12.017.

[17] P. Ayu, D. Pangastuti, ; Sukirno, and R. Efendi, “The Effect of Work Motivation and Compensation on Employee Performance,” Int. J. Multicult. Multireligious Underst., vol. 7, no. 3, pp. 292–299, 2020, [Online]. Available: http://ijmmu.com http://dx.doi//.org/10.18415/ijmmu.v7i3.1534%0Ahttp://dx.doi.org/10.18415/ijmmu.v7i3.1534

[18] R. Dhyan Parashakti, M. Ekhsan, and U. Dian Nusantara, “The Effect of Discipline and Motivation on Employee Performance in PT Samsung Elektronik Indonesia,” 2020, [Online]. Available: http://e-journal.stie-kusumanegara.ac.id

